# Anti-integrin αvβ6 antibodies predict pouchitis in patients with ulcerative colitis after restorative proctocolectomy with ileal pouch-anal anastomosis

**DOI:** 10.1101/2024.07.07.24309941

**Authors:** Risa Nakanishi, Takeshi Kuwada, Masahiro Shiokawa, Yoshihiro Nishikawa, Sakiko Ota, Hajime Yamazaki, Takafumi Yanaidani, Kenji Sawada, Ayako Hirata, Muneji Yasuda, Ikuhisa Takimoto, Koki Chikugo, Masataka Yokode, Yuya Muramoto, Shimpei Matsumoto, Tomoaki Matsumori, Norimitsu Uza, Tsutomu Chiba, Hiroshi Seno

## Abstract

**Background:** Pouchitis is the most common complication of restorative proctocolectomy (RPC) with ileal pouch-anal anastomosis (IPAA) in patients with ulcerative colitis (UC). We previously reported the presence of anti-integrin αvβ6 antibodies in the serum of patients with UC. This study investigated the association between anti-integrin αvβ6 antibodies and the development of pouchitis in patients with UC.

**Methods:** Serum levels of anti-integrin αvβ6 antibodies were measured by enzyme-linked immunosorbent assay in 16 patients with UC who underwent RPC with IPAA. Integrin αvβ6 expression in the colonic, terminal ileal, and pouch epithelium was examined using immunohistochemistry and western blot analysis.

**Results:** Anti-integrin αvβ6 antibody levels in patients with UC were significantly decreased at 3, 9, and 12 months after RPC (P < 0.05). However, in patients who developed pouchitis, antibody levels remained high. The antibody levels at the time of RPC were significantly higher in patients who developed pouchitis compared to those who did not. Kaplan–Meier analysis revealed a significantly higher incidence of pouchitis in patients with antibody levels above the cutoff at the time of RPC. Although integrin αvβ6 was not expressed in the terminal ileal epithelium at the time of RPC, expression became positive in the pouch epithelium of patients with pouchitis.

**Conclusions:** The anti-integrin αvβ6 antibody levels in patients with UC were decreased after RPC, but remained high in patients who developed pouchitis. The antibody levels at the time of RPC may serve as a potential prognostic biomarker for predicting the risk of pouchitis in patients with UC.

## Introduction

Ulcerative colitis (UC) is an inflammatory bowel disease characterized by chronic colonic mucosal inflammation. Despite medical advances, approximately 10–30% of patients with UC eventually require surgical intervention, such as restorative proctocolectomy (RPC) with ileal pouch-anal anastomosis (IPAA), because of medically refractory disease or the development of colitis-associated cancer. Recent studies have reported that the cumulative colectomy rates in patients with UC are 7.0% and 9.6% at 5 and 10 years after diagnosis, respectively [1]. Although these surgeries alleviate various UC-related symptoms, patients often develop pouchitis as a postoperative complication.

Pouchitis, the most common complication of RPC with IPAA, is characterized by inflammation of the ileal pouch, resembling UC. The incidence of pouchitis in patients with UC is 48% within the first year after IPAA, and almost 80% throughout the follow-up period after IPAA [2–4]. Interestingly, the incidence of pouchitis in patients with familial adenomatous polyposis, another condition that can be treated with IPAA, is remarkably lower at only 6% [5]. Therefore, immunological abnormalities may play important roles in the development of pouchitis in patients with UC. The endoscopic findings of pouchitis, such as mucosal redness, edema, erosions, and ulcers, resemble those observed in patients with UC. Moreover, treatments for pouchitis, including 5-aminosalicylic acid and corticosteroids, are similar to those for UC. Although the exact mechanism underlying pouchitis remains unknown, these similarities suggest a shared pathophysiology with UC.

We have previously identified anti-integrin αvβ6 antibodies in the serum of patients with UC [6]. Integrin αvβ6, a member of the integrin family, is expressed in the colonic epithelium and plays a crucial role in maintaining the epithelial barrier function through its interaction with extracellular matrix proteins, such as fibronectin [6–8]. Inhibition of integrin αvβ6–fibronectin binding by anti-integrin αvβ6 antibodies may lead to epithelial barrier dysfunction and contribute to the development of UC.

Considering the similarities in clinical presentation and treatment response between UC and pouchitis, and the potential role of anti-integrin αvβ6 antibodies in the pathophysiology of UC, we hypothesized that anti-integrin αvβ6 antibodies might also be involved in the development of pouchitis. This study aimed to investigate the association between serum anti-integrin αvβ6 antibody levels and the development of pouchitis in patients with UC who underwent RPC with IPAA.

## Materials and Methods

### Patients

We enrolled 16 patients with UC who underwent RPC with IPAA between September 2017 and January 2023 at Kyoto University Hospital (Table 1, Supplementary Table S1) and assessed the development of pouchitis until December 2023 (six patients with pouchitis and 10 patients without pouchitis). Patients were diagnosed based on symptoms, endoscopic, and histological findings, and the absence of alternative diagnoses [9, 10]. Pouchitis was defined as a modified Pouchitis Disease Activity Index of ≥ 5 [11, 12]. Serum samples were collected from September 2017 to December 2023 and stored at -80 °C until assayed. This study was performed in accordance with the Declaration of Helsinki and approved by the Ethics Committee of Kyoto University Graduate School and Faculty of Medicine (protocol number, R1004). Informed consent was obtained under the form of an opt-out on the website.

**Table 1.**
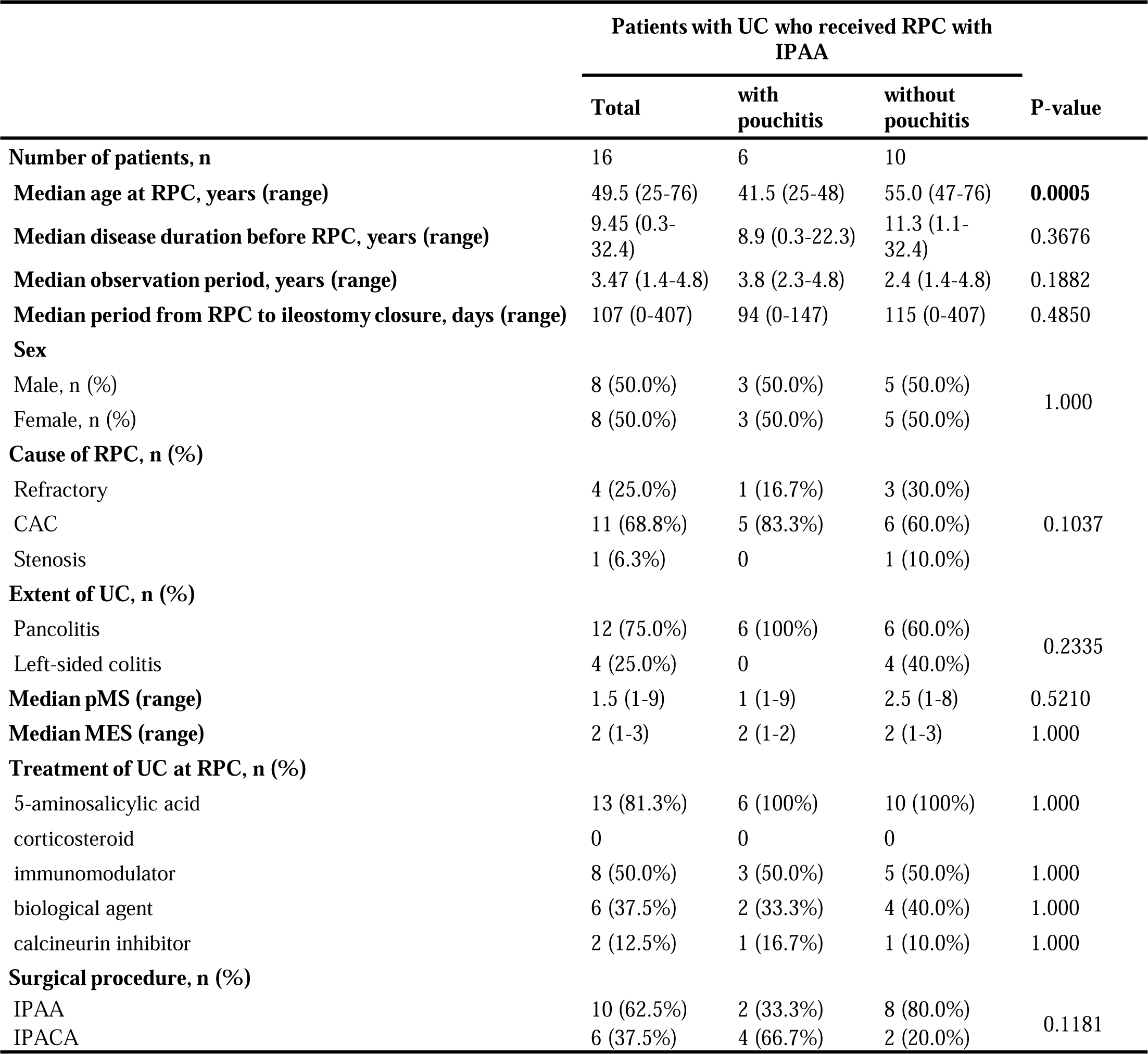
Clinical information of patients with UC who received RPC with IPAA, comparing between patients with and without pouchitis. Abbreviations: UC, ulcerative colitis; RPC, restorative proctocolectomy; CAC, colitis-associated cancer; pMS, partial Mayo score; MES, Mayo endoscopic subscore; IPAA, ileal pouch-anal anastomosis; IPACA, ileal pouch-anal canal anastomosis. Statistical analysis was performed using Mann–Whitney U test for continuous variables, and Fisher’s exact test for categorical variables.

### Enzyme-linked immunosorbent assay

To examine the serum anti-integrin αvβ6 antibody levels, we used Anti-Integrin αvβ6 ELISA Kit (5288, Medical and Biological Laboratories, Tokyo, Japan) according to the manufacturer’s instructions, with minor modifications. The standard material (recombinant monoclonal human anti-integrin αvβ6 antibody) was serially diluted with the reaction buffer from 400.0 U/mL to 0.781 U/mL. The reaction buffer served as the zero standard. The αvβ6-coated microplates were incubated with 100 µL of diluted serum (1:100) from patients or standard material in each well for 60 minutes at room temperature. After washing with wash solution, the plates were incubated with 100 µL of horseradish peroxidase-conjugated antibody at room temperature for 60 minutes. After another washing with wash solution, the bound reactants were detected by incubation for 20 minutes with 100 µL of substrate reagent. The absorbance was measured at the dual wavelengths of 450 and 620 nm after adding 100 µL of stop solution. The antibody levels were determined using a calibration curve generated based on the OD value of standard materials and blanks through a sigmoidal four-parameter logistic equation. When the antibody levels were higher than 400 U/mL, we retested with further diluted serums to ensure that the antibody levels of the diluted serums were less than 400 U/mL.

### Preparation of human immunoglobulin G

We used Ab-Rapid SPiN EX (P-014-10, ProteNova, Higashikagawa, Japan), according to the manufacturer’s instructions, to purify immunoglobulin (Ig) G from the serum of patients with UC. Purified IgG was stored at -20 °C [6, 13].

### Solid-phase integrin αvβ6 binding assay

A solid-phase integrin αvβ6 binding assay was performed according to the previously described method, with minor modifications, using ELISA Starter Accessory Kit (E101, Bethyl Laboratories, Montgomery, TX, USA) [6, 14–16]. A 96-well microlevel plate was coated with 100 µL/well of 2 µL/mL fibronectin (F0895, Sigma-Aldrich, St Louis, MO, USA) overnight at 4°C and blocked for 30 minutes at room temperature. Subsequently, 100 µL of diluted patient or control IgG (1:20) solution premixed with 2 µg/mL His-tagged integrin αvβ6 (IT6-H52E1, ACROBiosystems, Newark, NJ, USA) at 4°C overnight was added and incubated for 60 minutes at room temperature. After washing five times with the wash solution, 100 µL of peroxidase-conjugated anti-His-tag monoclonal antibody (1:5000, D291-7, Medical and Biological Laboratories) was added, followed by incubation at room temperature for 60 minutes. After another washing five times with the wash solution, bound reactants were detected by incubation with 3,3′,5,5′-tetramethylbenzidine for 10 minutes. The absorbance was measured at 450 nm. The assays were performed in the presence of MgCl_2_ and CaCl_2_ (1 mM each).

To calculate the inhibition rate, we used control wells coated with integrin αvβ6 and incubated with fibronectin in the absence of patient or control IgG. The inhibition rate was calculated using the following formula: (control optical density [OD]-sample OD) / control OD.

### Immunohistochemical analysis

Immunohistochemical analysis was performed using proximal colonic, terminal ileal, and ileal pouch tissues of patients with UC and patients with colorectal cancer (CRC) as controls, according to standard methods for human formalin-fixed paraffin-embedded samples. The primary antibody was anti-integrin β6 (1:3,000; NBP2-14136, Novus Biologicals, Centennial, CO, USA) and the secondary antibody was peroxidase-conjugated anti-rabbit IgG (K4003, Dako, Santa Clara, CA, USA). We enrolled five patients with CRC who underwent colectomy between October and December 2022 at Kyoto University Hospital (Supplementary Table S2).

### Western blot analysis

Protein extracts from human proximal colonic and terminal ileal mucosal samples of patients with UC and CRC were boiled in Laemmli sample buffer with 2.5% mercaptoethanol, fractionated on 4–15% sodium dodecyl sulfate polyacrylamide gels (4561086, Bio-Rad Laboratories, Hercules, CA, USA), and transferred to nitrocellulose membranes (10600003, Cytiva, Marlborough, MA, USA) according to standard protocols. Blocking and antibody reactions were performed using iBind Flex Western System (SLF2000, Thermo Fisher Scientific, Waltham, MA, USA) for 2.5 hours at room temperature, following the manufacturer’s protocol. The primary antibodies were anti-integrin αv (1:5,000; ab179475, Abcam, Cambridge, UK), anti-integrin β6 (1:10,000; ab187155, Abcam), and anti-β actin (1:10,000; ab6276, Abcam). The secondary antibodies were peroxidase-conjugated anti-mouse IgG (1:10,000; A28177, Thermo Fisher Scientific) and anti-rabbit IgG (1:10,000; 31458, Thermo Fisher Scientific). Protein-antibody complexes were visualized using a chemiluminescent substrate (34095, Thermo Fisher Scientific). Immunoreactive protein bands were detected using Amersham Imager 600 (Cytiva). As controls, we enrolled five patients with CRC (Supplementary Table S2).

### Statistical analysis

Statistical analysis was performed using GraphPad Prism Version 10 (GraphPad Software, San Diego, CA, USA). For all boxplots, the box represents the interquartile range, the centerline represents the median, and the whiskers indicate the minimum to maximum value.

Serum anti-integrin αvβ6 antibody levels at 0, 3, 6, 9, and 12 months after RPC and the inhibition of integrin αvβ6–fibronectin binding at one year after RPC were compared between patients who developed pouchitis and those who did not using Mann–Whitney U test. Changes in serum anti-integrin αvβ6 antibody levels over time (0, 3, 6, 9, and 12 months after RPC) were compared using Wilcoxon matched-pairs signed-rank test. The association between antibody levels and future development of pouchitis was assessed using receiver operating characteristic curve analysis. The optimal cutoff value was determined by the Youden index. Kaplan–Meier analysis was performed to compare the pouchitis-free survival probability between patients with antibody levels above and below the cutoff value at the time of RPC by the log-rank test. Patients were censored at the time of last follow-up or study end date (December 2023). The correlation between anti-integrin αvβ6 antibody levels and the inhibition of integrin αvβ6–fibronectin binding was evaluated by means of the Pearson product moment correlation. Differences were considered statistically significant at P < 0.05.

## Results

### Anti-integrin αvβ6 antibody levels in patients with ulcerative colitis after restorative proctocolectomy

We examined the serum anti-integrin αvβ6 antibody levels of 16 patients with UC who underwent RPC with IPAA by enzyme-linked immunosorbent assay. Anti-integrin αvβ6 antibody levels of patients with UC significantly decreased after RPC (P < 0.05, Fig. 1a). However, in patients who developed pouchitis, the antibody levels remained high even after RPC (Fig. 1b). Subsequently, we compared the anti-integrin αvβ6 antibody levels at the time of RPC in patients who developed pouchitis and those who did not. The anti-integrin αvβ6 antibody levels in patients who developed pouchitis were significantly higher than those in patients who did not (P < 0.05, Fig. 1c, Fig. 2a). Table 1 shows the clinical characteristics of patients who developed pouchitis and those who did not. Patients who developed pouchitis were significantly younger at the time of RPC compared to those who did not (median age: 41.5 years versus 55.0 years, P = 0.0005).

**Fig. 1.**
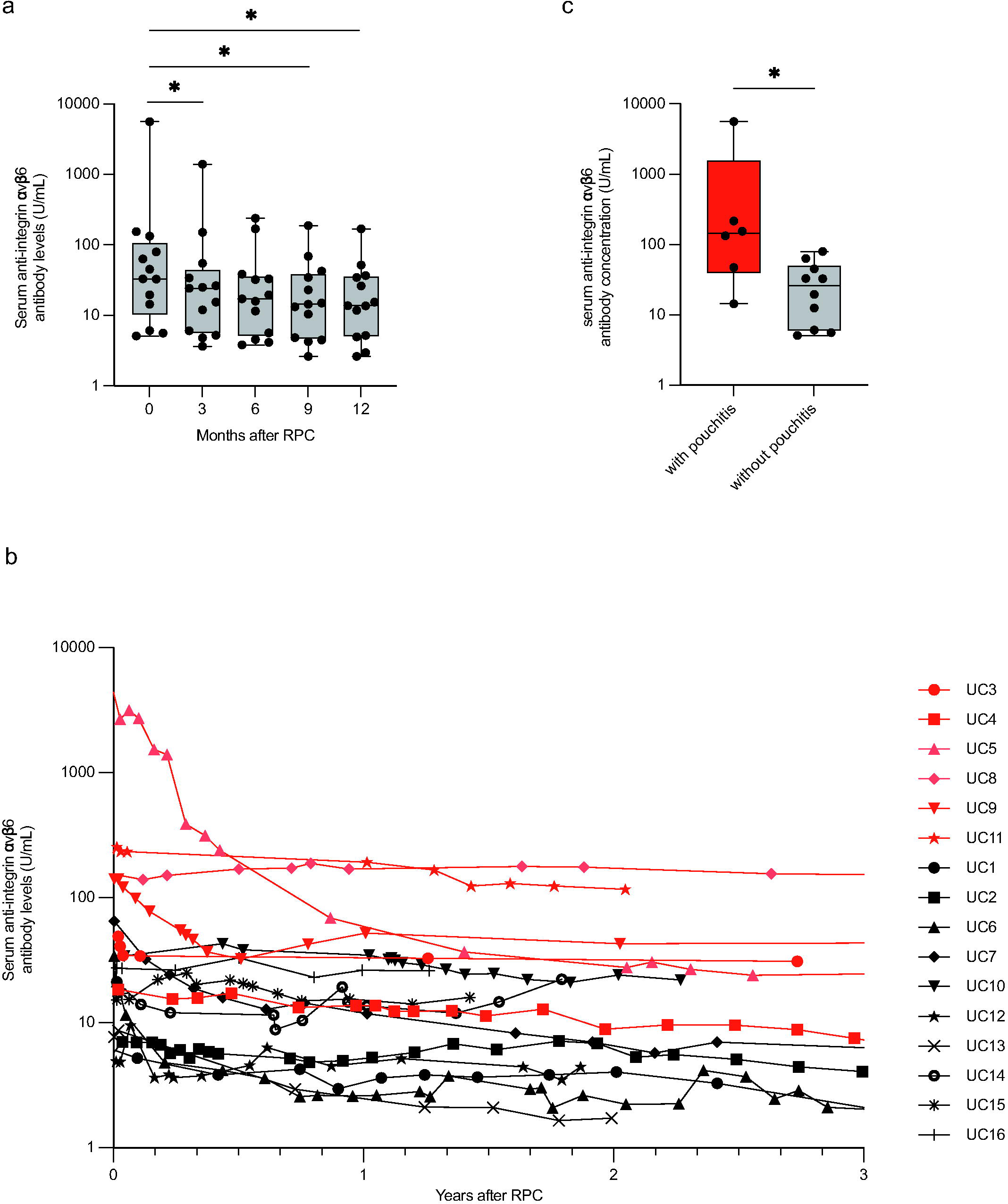
A**n**ti**-integrin** α**v**β**6 antibody levels in serum samples of patients with ulcerative colitis (UC) who received restorative proctocolectomy (RPC) with ileal pouch-anal anastomosis** Serum immunoglobulin (Ig) G antibodies against integrin αvβ6 were quantified by enzyme-linked immunosorbent assay. (a) Changes of serum anti-integrin αvβ6 antibody levels in 13 patients with UC after RPC. The anti-integrin αvβ6 antibody levels at 3, 9, and 12 months after RPC were significantly lower than those at the time of RPC (three out of 16 patients were excluded from the analysis as serum samples at 3, 6, and 9 months after RPC were not available). *P < 0.05, Wilcoxon matched-pairs signed-rank test. (b) Overall change of the anti-integrin αvβ6 antibody levels in patients with (red) and without (black) pouchitis after RPC (n = 16). The anti-integrin αvβ6 antibody levels in patients who developed pouchitis remained high. (c) Comparison of the anti-integrin αvβ6 antibody levels at the time of RPC between patients who developed pouchitis and those who did not. The antibody levels in patients who developed pouchitis were significantly higher than those in patients who did not develop pouchitis at the time of RPC (n = 16). *P < 0.05, Mann–Whitney U test.

**Fig. 2.**
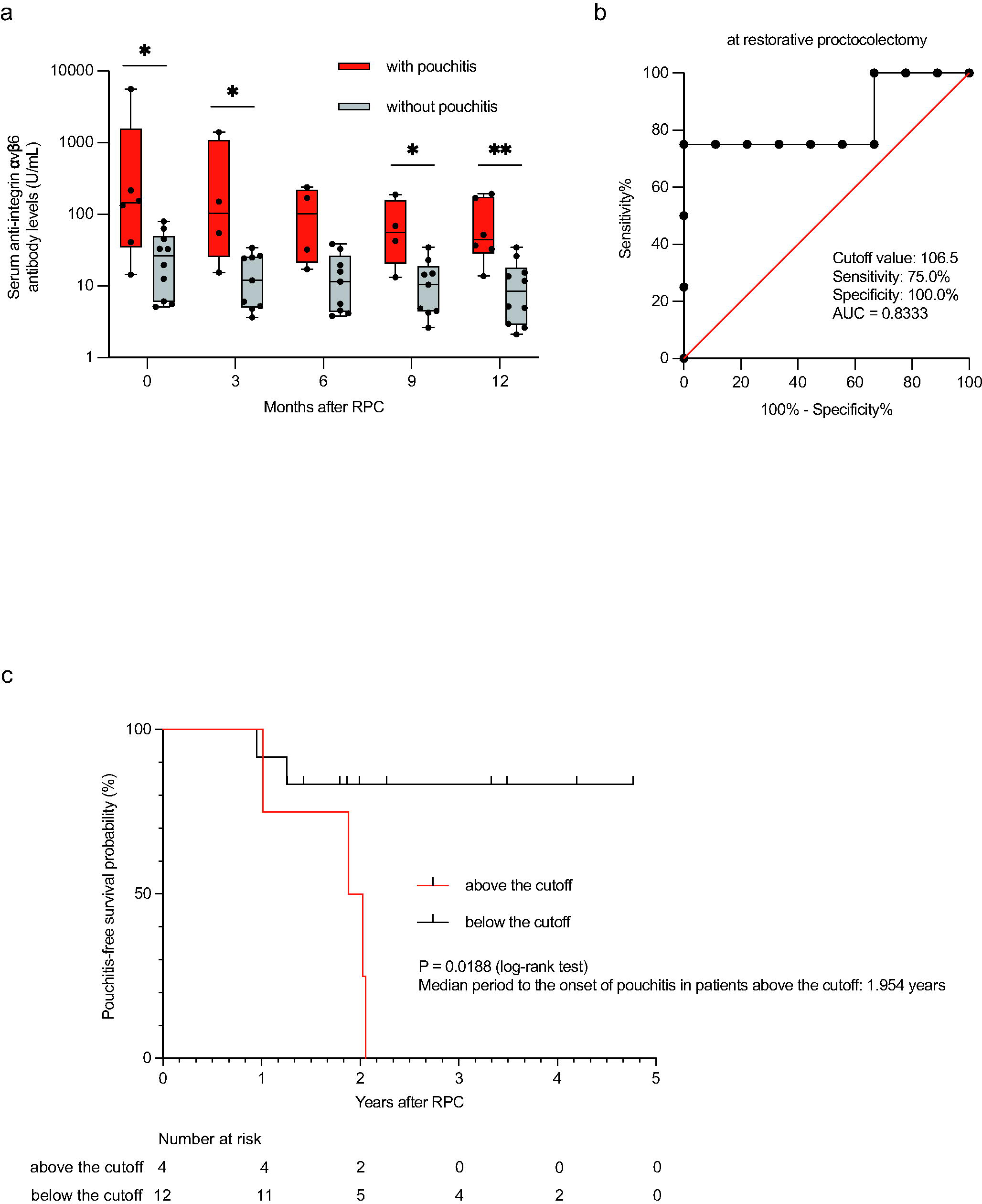
Prediction of the risk of pouchitis based on anti-integrin **α**v**β**6 antibody levels at the time of RPC (a) Serial changes of anti-integrin αvβ6 antibody levels at 0, 3, 6, 9, and 12 months after RPC between patients who developed pouchitis and those who did not (n = 16, the serum samples of three out of 16 patients at 3, 6, and 9 months after RPC were not available). At 0, 3, 9, and 12 months after RPC, the anti-integrin αvβ6 antibody levels in patients who developed pouchitis were significantly higher than those in patients who did not. *P < 0.05, **P < 0.01, Mann–Whitney U test. (b) Receiver operating characteristic (ROC) curve of serum anti-integrin αvβ6 antibody levels at the time of RPC (n = 13). Area under the curve (AUC) = 0.8333, cutoff value = 106.5 U/mL. The sensitivity and specificity for the onset of pouchitis were 75% and 100%, respectively. (c) Kaplan–Meier analysis for pouchitis-free survival probability after RPC. Patients with anti-integrin αvβ6 antibody levels above the cutoff value and those below the cutoff value were compared (n = 16). P = 0.0188, log-rank test. The median period from RPC to the onset of pouchitis in patients with anti-integrin αvβ6 antibody levels above the cutoff was 1.954 years.

### Prediction of the risk of pouchitis based on anti-integrin αvβ6 antibody levels

We compared serial changes of the anti-integrin αvβ6 antibody levels at 0, 3, 6, 9, and 12 months after RPC between patients who developed pouchitis and those who did not. In addition to the antibody levels at the time of RPC, significant differences were observed in anti-integrin αvβ6 antibody levels at 3, 9, and 12 months after RPC (Fig. 2a). To establish a predictive marker for the risk of pouchitis, we determined the optimal cutoff value for anti-integrin αvβ6 antibody levels at the time of RPC through receiver operating characteristic curve analysis, which was found to be 106.5 U/mL (Fig. 2b, Supplementary Fig. S1). The sensitivity and specificity of this predictive marker for the onset of pouchitis were 75% and 100%, respectively. Patients with anti-integrin αvβ6 antibody levels above the cutoff had a significantly higher incidence of pouchitis than those below the cutoff, as demonstrated by Kaplan–Meier analysis (Fig. 2c). The median period from RPC to the onset of pouchitis in patients with anti-integrin αvβ6 antibody levels above the cutoff was 1.95 years. These findings suggest the usefulness of anti-integrin αvβ6 antibody levels at the time of RPC as a predictive marker for development of pouchitis.

### Inhibition of integrin αvβ6–fibronectin binding by IgG from patients with pouchitis

We previously reported that anti-integrin αvβ6 antibodies in patients with UC inhibit integrin αvβ6–fibronectin binding [6]. Therefore, we examined whether anti-integrin αvβ6 antibodies in patients with pouchitis also have an inhibitory activity on integrin αvβ6–fibronectin binding. Using purified IgG from patients with UC at one year after RPC, we conducted solid-phase integrin αvβ6–fibronectin binding assays. The results revealed that IgG from patients who developed pouchitis effectively blocked integrin αvβ6–fibronectin binding (Fig. 3a, b). Importantly, the inhibitory activity of IgG correlated with the levels of anti-integrin αvβ6 antibodies, as in patients with UC [6].

**Fig. 3.**
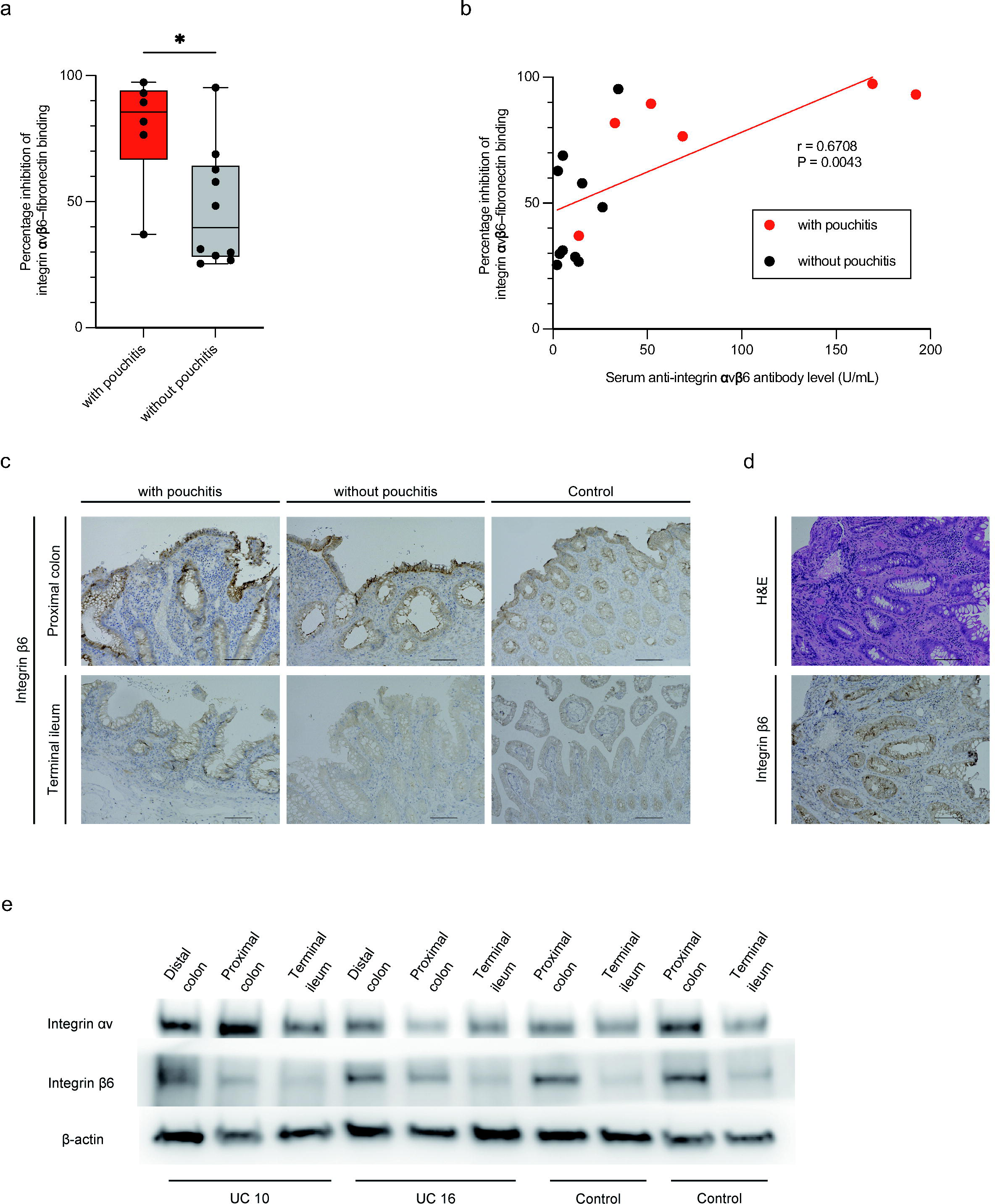
Inhibition of integrin **α**v**β**6**–**fibronectin binding by IgG extracted from patients with UC and expression of integrin **α**v**β**6 in proximal colonic, terminal ileal, and ileal pouch epithelium (a) The percentage inhibition of integrin αvβ6–fibronectin binding by serum IgG in patients who developed pouchitis was significantly higher than that in patients who did not develop pouchitis at one year after RPC (n = 16). *P < 0.05, Mann–Whitney U test. (b) Relationship between the percentage inhibition of integrin αvβ6–fibronectin binding by serum IgG and anti-integrin αvβ6 antibody levels in patients who developed pouchitis and those who did not at one year after RPC (n = 16). The inhibitory activity of IgG samples in patients with UC was correlated with the levels of anti-integrin αvβ6 antibodies. Pearson product moment correlation. r = 0.6708, P = 0.0043. (c) Representative immunohistochemical staining with anti-integrin β6 antibody in proximal colonic and terminal ileal epithelium surgical samples of three patients with UC who developed pouchitis and four patients who did not at the time of RPC, and five patients with colorectal cancer as controls at the time of colectomy. Scale bars, 100 µm. (d) Representative hematoxylin and eosin staining and immunohistochemical staining of ileal pouch epithelium biopsy specimen of five patients with pouchitis at pouch endoscopy with anti-integrin β6 antibody. Scale bars, 100 µm. (e) Representative western blot analysis with anti-integrin αv and β6 antibodies of proximal colonic and terminal ileal mucosal frozen samples in six patients with UC at the time of RPC, and in five patients with colorectal cancer as controls at the time of colectomy.

### Expression of integrin αvβ6 in proximal colonic, terminal ileal, and ileal pouch epithelium

Colonic metaplasia of the ileal pouch has been proposed as one of the causes of pouchitis [17, 18]. Integrin αvβ6 is known to be expressed in the colonic epithelium, but its expression in the terminal ileal epithelium remains unclear [6,7]. To examine the expression of integrin αvβ6 in the proximal colonic, terminal ileal, and ileal pouch epithelium, we conducted immunohistochemical analysis. At the time of RPC, integrin αvβ6 was expressed in the proximal colonic epithelium, but not in the terminal ileal epithelium in all patients with UC that were examined (Fig. 3c). Interestingly, however, integrin αvβ6 expression could be observed in the ileal pouch epithelium of the patients who developed pouchitis (Fig. 3d). In addition, we performed western blot analysis to quantitatively evaluate integrin αvβ6 expression at the time of RPC. The results showed that integrin αvβ6 was highly expressed in the proximal colonic mucosa of patients with UC compared to in the mucosa of non-UC controls (patients with CRC), but it was barely expressed in the terminal ileal mucosa (Fig. 3e).

## Discussion

We previously reported the presence of anti-integrin αvβ6 antibodies in patients with UC with high sensitivity and specificity. In this study, we found that anti-integrin αvβ6 antibody levels in patients with UC significantly decreased after RPC, however, the antibody levels in patients who developed pouchitis remained high, even one year after RPC. Interestingly, although integrin αvβ6 was not expressed in the terminal ileal mucosa of patients with UC, expression became positive in the ileal pouch epithelium of the patients who developed pouchitis. Most importantly, preoperative anti-integrin αvβ6 antibody levels in patients who developed pouchitis were significantly higher than those in patients who did not develop pouchitis. These data suggest that anti-integrin αvβ6 antibody levels may be a useful marker for predicting the development of pouchitis. Previously, we reported that IgG in patients with UC inhibit integrin αvβ6–fibronectin binding [6]. In this study, we confirmed that postoperative IgG levels in patients with UC have similar inhibitory effects.

To date, the precise mechanism for the decrease of anti-integrin αvβ6 antibodies after RPC in patients with UC remains unknown. However, integrin αvβ6 is expressed in the colonic epithelium, and its expression is increased in inflammatory conditions [6,7]. Thus, removal of the inflammatory colon with resulting disappearance of integrin αvβ6 as the autoantigen may have resulted in the decrease of anti-integrin αvβ6 antibody levels after RPC.

However, it should be emphasized that, when focusing on the patients who developed pouchitis, the antibody levels remained high even after RPC with IPAA. In this regard, it is noteworthy that, although integrin αvβ6 was not expressed in the ileal mucosa of patients with UC at the time of RPC, expression became positive in the ileal pouch mucosa of the patients who developed pouchitis in this study. Previous reports showing that the ileal pouch epithelium undergoes colonic metaplasia following pouch formation in patients with UC offer consistent results with the present data [17, 18]. Taken together, the results suggested that expression of integrin αvβ6 in the ileal pouch mucosa after RPC with IPAA may continuously stimulate production of anti-integrin αvβ6 antibody.

In previous studies, we found that anti-integrin αvβ6 antibodies in patients with UC had inhibitory effects on integrin αvβ6–fibronectin binding, and showed a positive correlation between anti-integrin αvβ6 antibody levels and disease activity, suggesting involvement of the antibody in the pathophysiology of UC [6]. In this study, we observed that postoperative patients’ serum also had inhibitory effects on integrin αvβ6–fibronectin binding, and these effects correlated with the levels of anti-integrin αvβ6 antibodies, similar to the findings in patients with UC before RPC. Thus, it may be possible that anti-integrin αvβ6 antibodies are involved in the development of pouchitis by disrupting the epithelial barrier function through inhibition of integrin αvβ6–fibronectin binding. Taken together, these findings suggest that antibody production induced by integrin αvβ6 expressed in the pouch mucosa, and the inhibition of integrin αvβ6–fibronectin binding by anti-integrin αvβ6 antibodies together might create a vicious cycle in the development of pouchitis.

A recent study has shown that anti-integrin αvβ6 antibodies could be detected up to 10 years before the diagnosis of UC, indicating that the antibodies can be a biomarker for predicting the disease onset [19]. Therefore, we investigated the usefulness of anti-integrin αvβ6 antibodies as a biomarker for predicting the development of pouchitis following RPC with IPAA. First, we found that anti-integrin αvβ6 antibody levels before RPC in patients who developed pouchitis were significantly higher than those who did not. Moreover, using the optimal cutoff value of anti-integrin αvβ6 antibody levels determined through receiver operation characteristic analysis at the time of RPC, the sensitivity and the specificity of the antibody levels for predicting the development of pouchitis were 75% and 100%, respectively. Furthermore, Kaplan–Meier analysis demonstrated a significantly higher incidence of pouchitis in patients with antibody levels above the cutoff. These data strongly suggested that anti-integrin αvβ6 antibodies can be a novel and excellent biomarker for predicting the development of pouchitis after RPC with IPAA in patients with UC, and that measuring the anti-integrin αvβ6 antibody levels enables early detection and therapeutic intervention contributing to the prevention of pouchitis.

The limitation of this study was the small number of patients from a single institution. Further research involving larger patient cohorts from multiple institutions is required to validate these results.

In conclusion, anti-integrin αvβ6 antibody levels in patients with UC were significantly decreased after RPC. However, the antibody levels remained high in patients who developed pouchitis. Since anti-integrin αvβ6 antibody levels before RPC in patients who developed pouchitis were significantly higher than those who did not, the antibody levels may be useful for predicting development of pouchitis after RPC with IPAA in patients with UC.

## Data Availability

All data produced in the present work are contained in the manuscript

## Acknowledgments

We thank the patients who provided serum samples for this study. We would also like to thank Shino Yamaguchi and Taichi Ito for their valuable technical support. We are grateful to Medical and Biological Laboratories Co., Ltd. for providing the Anti-Integrin αvβ6 ELISA Kit used in this study. Finally, we thank Editage for English language editing.

## Author contributions

Conceptualization: Risa Nakanishi, Takeshi Kuwada, Masahiro Shiokawa, and Yoshihiro Nishikawa; Methodology: Risa Nakanishi and Takeshi Kuwada; Formal analysis: Risa Nakanishi and Hajime Yamazaki; Investigation: Risa Nakanishi and Takeshi Kuwada; Funding acquisition: Takeshi Kuwada; Resources: Risa Nakanishi, Takeshi Kuwada, and Sakiko Ota; Supervision: Tsutomu Chiba and Hiroshi Seno; Visualization: Risa Nakanishi; Writing - original draft preparation: Risa Nakanishi and Takeshi Kuwada; Writing - review and editing: Masahiro Shiokawa, Yoshihiro Nishikawa, Sakiko Ota, Hajime Yamazaki, Takafumi Yanaidani, Kenji Sawada, Ayako Hirata, Muneji Yasuda, Ikuhisa Takimoto, Koki Chikugo, Masataka Yokode, Yuya Muramoto, Shimpei Matsumoto, Tomoaki Matsumori, Norimitsu Uza, Tsutomu Chiba, and Hiroshi Seno.

## Supplementary information

Supplementary Table S1. Clinical information of patients with UC

Abbreviations: UC, ulcerative colitis; CAC, colitis-associated cancer; pMS, partial Mayo score; MES, Mayo endoscopic subscore; mPDAI, modified Pouchitis Disease Activity Index; F, female; M, male; IPAA, ileal pouch-anal anastomosis; IPACA, ileal pouch-anal canal anastomosis.

Supplementary Table S2. Clinical information of patients with CRC

Abbreviations: CRC, colorectal cancer; M, male; F, female

Supplementary Fig. S1 ROC curve of serum anti-integrin αvβ6 antibody levels at 3, 6, 9, and 12 months after RPC (n = 13) and Kaplan–Meier analysis for pouchitis-free survival probability after RPC (n = 16)

(a) ROC curve at 3 months after RPC. AUC = 0.8889; cutoff value = 44.50 U/mL. The sensitivity and specificity for the onset of pouchitis were 75% and 100%, respectively.

(b) Kaplan–Meier analysis of the pouchitis-free survival probability at 3 months after RPC. P = 0.0188, log-rank test. The median period from RPC to the onset of pouchitis in patients with anti-integrin αvβ6 antibody levels above the cutoff was 1.704 years from 3 months after RPC.

(c) ROC curve at 6 months after RPC. AUC = 0.8611; cutoff value = 13.0 U/mL. The sensitivity and specificity for the onset of pouchitis were 100% and 66.67%, respectively.

(d) Kaplan–Meier analysis for pouchitis-free survival probability at 6 months after RPC. P = 0.0376; log-rank test. The median period from RPC to the onset of pouchitis in patients with anti-integrin αvβ6 antibody levels above the cutoff was 1.382 years from 6 months after RPC.

(e) ROC curve at 9 months after RPC. AUC = 0.8889; cutoff value = 38.53 U/mL. The sensitivity and specificity for onset of pouchitis were 75% and 100%, respectively.

(f) Kaplan–Meier analysis for pouchitis-free survival probability at 9 months after RPC. P = 0.0188; log-rank test. The median period from RPC to the onset of pouchitis in patients with anti-integrin αvβ6 antibody levels above the cutoff was 1.204 years from 9 months after RPC.

(g) ROC curve at 12 months after RPC. AUC = 0.9167; cutoff value = 35.70 U/mL. The sensitivity and specificity for onset of pouchitis were 75% and 100%, respectively.

(h) Kaplan–Meier analysis for pouchitis-free survival probability at 12 months after RPC. P = 0.0046, log-rank test. The median period from RPC to the onset of pouchitis in patients with anti-integrin αvβ6 antibody levels above the cutoff was 0.9535 years from 12 months after RPC.

