## Supplementary figures and images for "Anti-integrin αvβ6 antibodies predict pouchitis in patients with ulcerative colitis after restorative proctocolectomy with ileal pouch-anal anastomosis"

### Supplementary Fig. S1

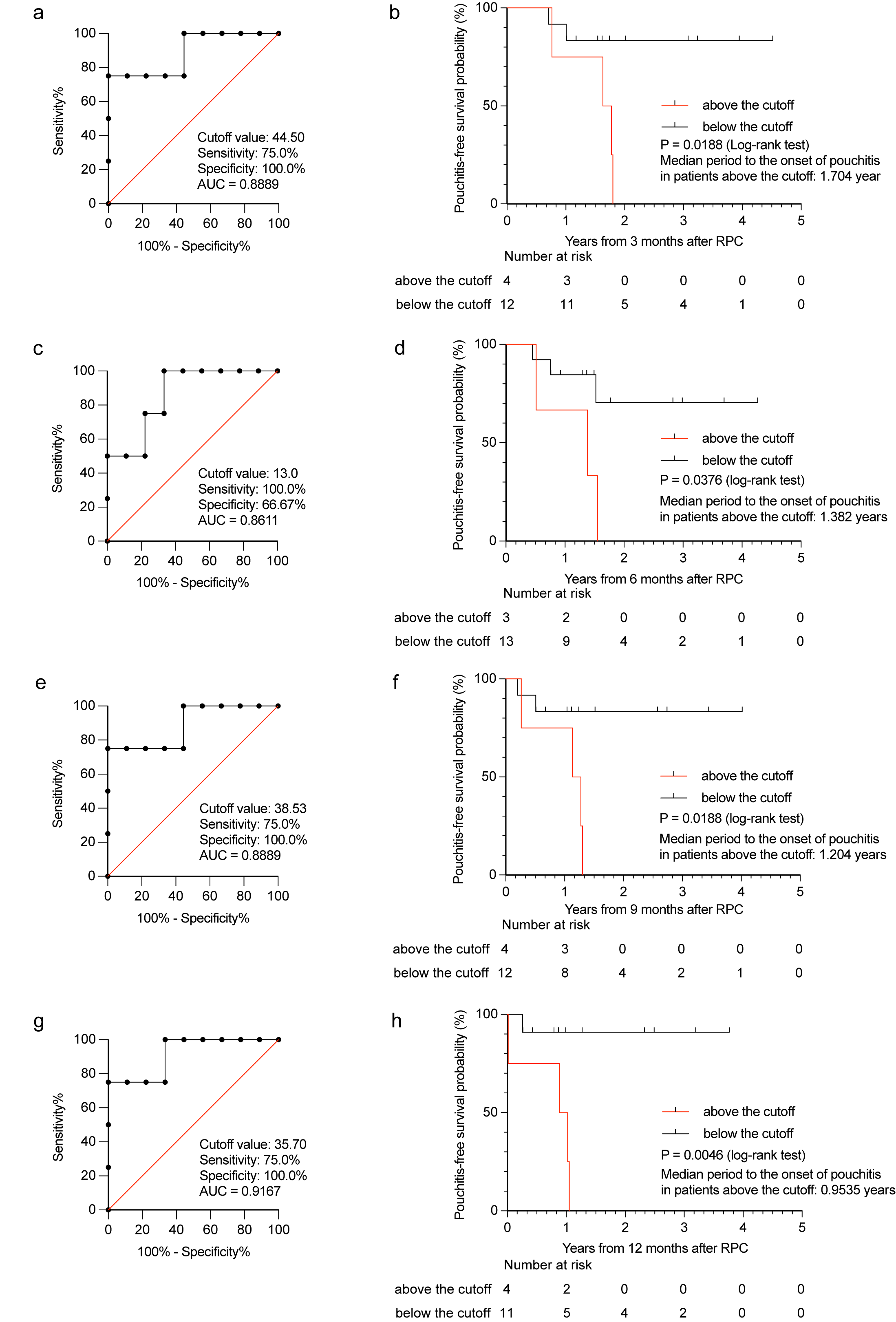
