## Supplementary Table S1 for "Anti-integrin αvβ6 antibodies predict pouchitis in patients with ulcerative colitis after restorative proctocolectomy with ileal pouch-anal anastomosis"

### Slide 1
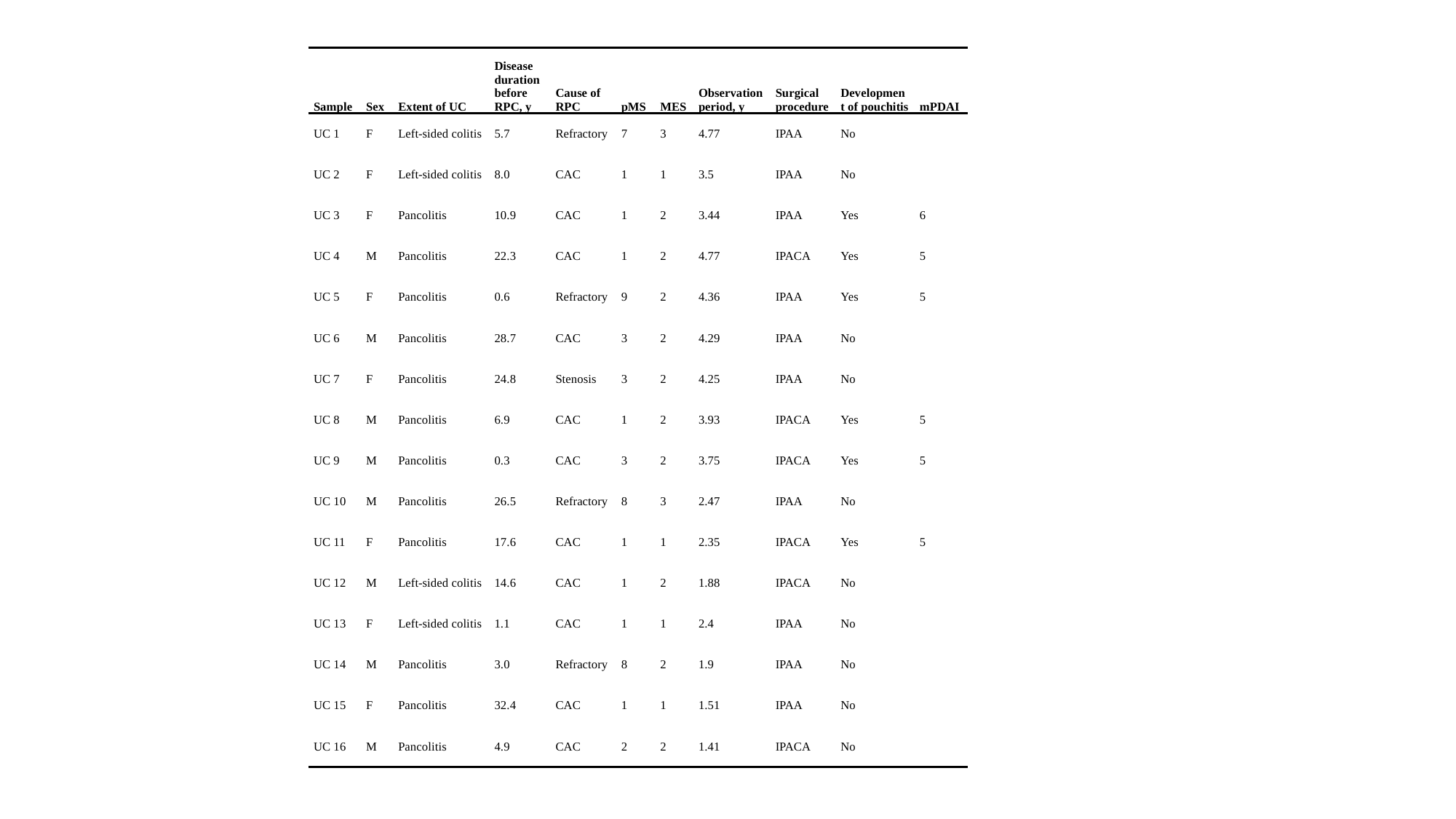

| Sample | Sex | Extent of UC | Disease duration before RPC, y | Cause of RPC | pMS | MES | Observation period, y | Surgical procedure | Development of pouchitis | mPDAI |
| --- | --- | --- | --- | --- | --- | --- | --- | --- | --- | --- |
| UC 1 | F | Left-sided colitis | 5.7 | Refractory | 7 | 3 | 4.77 | IPAA | No | |
| UC 2 | F | Left-sided colitis | 8.0 | CAC | 1 | 1 | 3.5 | IPAA | No | |
| UC 3 | F | Pancolitis | 10.9 | CAC | 1 | 2 | 3.44 | IPAA | Yes | 6 |
| UC 4 | M | Pancolitis | 22.3 | CAC | 1 | 2 | 4.77 | IPACA | Yes | 5 |
| UC 5 | F | Pancolitis | 0.6 | Refractory | 9 | 2 | 4.36 | IPAA | Yes | 5 |
| UC 6 | M | Pancolitis | 28.7 | CAC | 3 | 2 | 4.29 | IPAA | No | |
| UC 7 | F | Pancolitis | 24.8 | Stenosis | 3 | 2 | 4.25 | IPAA | No | |
| UC 8 | M | Pancolitis | 6.9 | CAC | 1 | 2 | 3.93 | IPACA | Yes | 5 |
| UC 9 | M | Pancolitis | 0.3 | CAC | 3 | 2 | 3.75 | IPACA | Yes | 5 |
| UC 10 | M | Pancolitis | 26.5 | Refractory | 8 | 3 | 2.47 | IPAA | No | |
| UC 11 | F | Pancolitis | 17.6 | CAC | 1 | 1 | 2.35 | IPACA | Yes | 5 |
| UC 12 | M | Left-sided colitis | 14.6 | CAC | 1 | 2 | 1.88 | IPACA | No | |
| UC 13 | F | Left-sided colitis | 1.1 | CAC | 1 | 1 | 2.4 | IPAA | No | |
| UC 14 | M | Pancolitis | 3.0 | Refractory | 8 | 2 | 1.9 | IPAA | No | |
| UC 15 | F | Pancolitis | 32.4 | CAC | 1 | 1 | 1.51 | IPAA | No | |
| UC 16 | M | Pancolitis | 4.9 | CAC | 2 | 2 | 1.41 | IPACA | No | |
