## Supplementary Table S2 for "Anti-integrin αvβ6 antibodies predict pouchitis in patients with ulcerative colitis after restorative proctocolectomy with ileal pouch-anal anastomosis"

### Slide 1
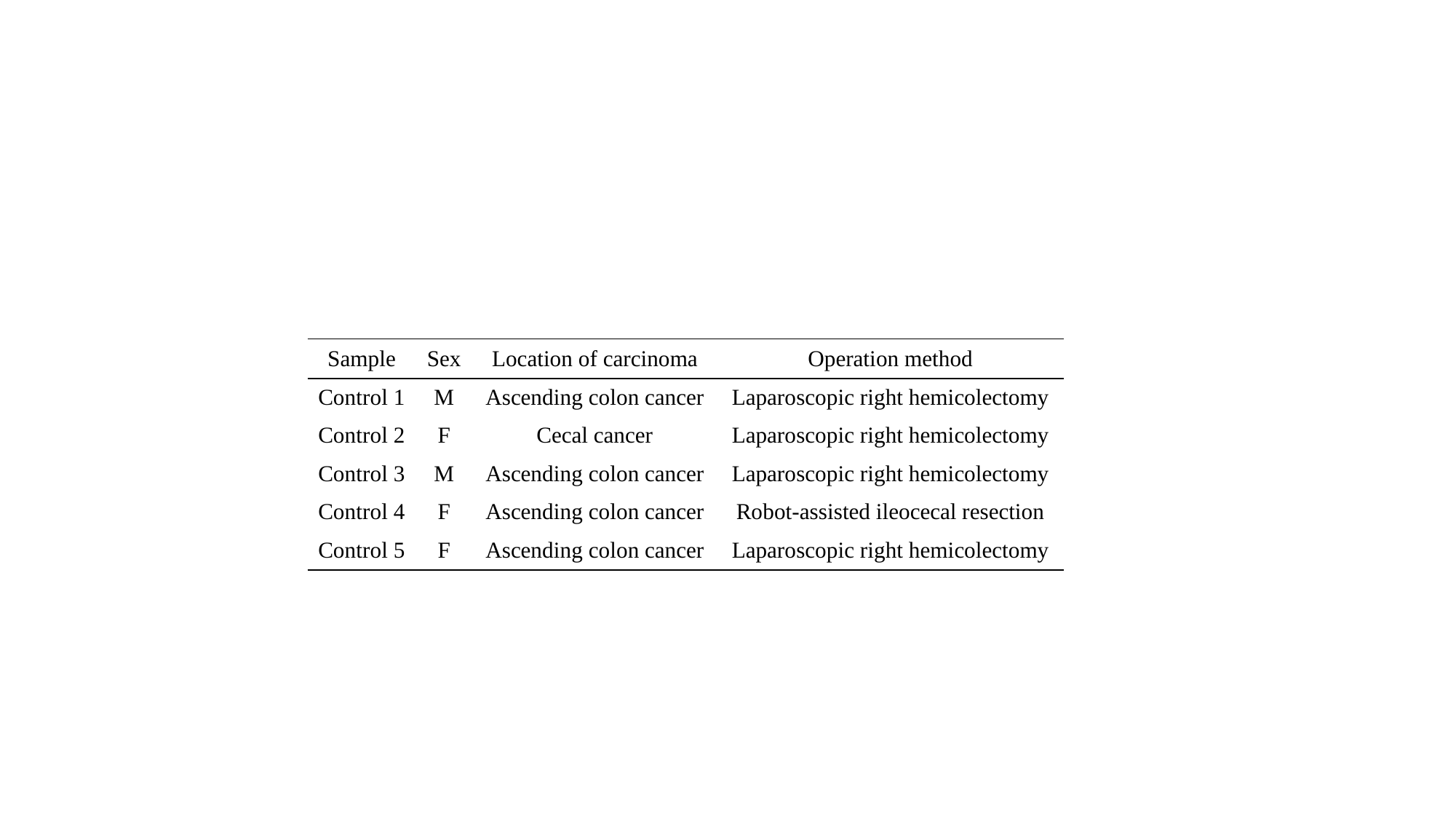

| Sample | Sex | Location of carcinoma | Operation method |
| --- | --- | --- | --- |
| Control 1 | M | Ascending colon cancer | Laparoscopic right hemicolectomy |
| Control 2 | F | Cecal cancer | Laparoscopic right hemicolectomy |
| Control 3 | M | Ascending colon cancer | Laparoscopic right hemicolectomy |
| Control 4 | F | Ascending colon cancer | Robot-assisted ileocecal resection |
| Control 5 | F | Ascending colon cancer | Laparoscopic right hemicolectomy |
